# Sequential Bilateral Accelerated Theta Burst Stimulation in Adolescents With Suicidal Ideation Associated With Major Depressive Disorder: Protocol for a Randomized Controlled Trial

**DOI:** 10.1101/2022.12.21.22283783

**Authors:** Deniz Yuruk, Can Ozger, Juan F. Garzon, Jarrod M. Leffler, Julia Shekunov, Jennifer L. Vande Voort, Michael J. Zaccariello, Paul A. Nakonezny, Paul E. Croarkin

**Affiliations:** Research Fellow in the Department of Psychiatry and Psychology; Mayo Clinic School of Graduate Medical Education, Mayo Clinic College of Medicine and Science, Rochester, Minnesota, Virginia Treatment Center for Children; Virginia Commonwealth University, Richmond, Virginia; Clinical Research Coordinator in the Department of Psychiatry and Psychology; Department of Psychiatry and Psychology; Mayo Clinic Children’s Research Center; Mayo Clinic Depression Center; Mayo Clinic, Rochester, Minnesota and Department of Population and Data Sciences; UT Southwestern Medical Center, Dallas, Texas.

**Keywords:** accelerated theta burst stimulation, adolescent, depression, neuromodulation, suicidal ideation, suicide, transcranial magnetic stimulation

## Abstract

**Background:** Suicide is a leading cause of death in adolescents worldwide. Previous research findings suggest that suicidal adolescents with depression have pathophysiological dorsolateral prefrontal cortex (DLPFC) deficits in γ-aminobutyric acid neurotransmission. Interventions with transcranial magnetic stimulation (TMS) directly address these underlying pathophysiological deficits in the prefrontal cortex. Theta burst stimulation (TBS) is newer dosing approach for TMS. Accelerated TBS (aTBS) involves administering multiple sessions of TMS daily as this dosing may be more efficient, tolerable, and rapid acting than standard TMS.

**Methods:** This is a randomized, double-blind, sham-controlled trial of sequential bilateral aTBS in adolescents with MDD and suicidal ideation. Three sessions are administered daily for 10 days. During each session, continuous TBS is administered first to the right DPFC, in which 1,800 pulses are delivered continuously over 120 seconds. Then intermittent TBS is applied to the left DPFC, in which 1,800 pulses are delivered in 2-second bursts and repeated every 10 seconds for 570 seconds. The TBS parameters were adopted from prior research, with 3-pulse, 50-Hz bursts given every 200 ms (at 5 Hz) with an intensity of 80% active motor threshold. The comparison group will receive 3 daily sessions of bilateral sham TBS treatment for 10 days. All participants will receive the standard of care for patients with depression and suicidal ideation including daily psychotherapeutic skill sessions. Long-interval intracortical inhibition (LICI) biomarkers will be measured before and after treatment. Exploratory measures will be collected with TMS and electroencephalography for biomarker development.

**Discussion:** This is the first known randomized controlled trial to examine the efficacy of sequential bilateral aTBS for treating suicidal ideation in adolescents with MDD. Results from this study will also provide opportunities to further understand the neurophysiological and molecular mechanisms of suicidal ideation in adolescents.

**Trial registration:** Investigational device exemption (IDE) Number: G200220, ClinicalTrials.gov (ID: NCT04701840). Registered August 6, 2020. https://clinicaltrials.gov/ct2/show/NCT04502758?term=NCT04701840&draw=2&rank=1

## Introduction

Suicide is a leading cause of death in adolescents worldwide [1–3]. In the US, 18% of teenagers report seriously considering suicide, 15% have made a suicide plan, and 9% have attempted suicide within the preceding year [4, 5]. Effective treatments of suicidal ideation are limited for adolescents with major depressive disorder (MDD) [6–8]. Previous research focused on understanding the risk factors for suicide in adolescents in attempted to reduce their overall suicide rates [4,8–10]. Despite decades of research, however, the predictive value of identified suicide risk factors is low and has not improved substantially [8]. Biological correlates explored to date are also weak predictors of suicidal behaviors [8,11–13]. Little prior research has focused on rapid-acting treatments for adolescents with MDD who are actively struggling with thoughts of suicide or have recently attempted suicide [7, 14]. Although some studies explored the antisuicidal effects of alternative treatments in adult populations, a considerable knowledge gap remains regarding treating adolescents at risk for suicide [8,15,16].

Suicidal persons have structural, functional, neurophysiological, and neuroplastic alterations in the dorsolateral prefrontal cortex (DPFC) [17–19], and these alterations may be targeted with neurostimulation. Structural alterations and serotonergic dysfunction in the DPFC are associated with lethality of suicide attempts [19–21]. Suicidal persons also have abnormalities in processing negative emotional stimuli and in activating the DPFC. These findings for DPFC activation vary across studies, suggesting multiple neural phenotypes of suicidality that are poorly understood [19]. Previous research findings suggest that suicidal adolescents with depression have pathophysiological DPFC deficits in γ-aminobutyric acid (GABA) function [22–25]. Impaired prefrontal cortical GABAergic inhibitory gating related to depression may reduce an adolescent’s control over their thoughts and behaviors and may culminate in suicidal ideation and behaviors [17,26,27]. Interventions with transcranial magnetic stimulation (TMS) directly address these underlying pathophysiological deficits in the DPFC [28, 29].

In the past decade, studies of TMS interventions found that bilateral TMS strongly reduced suicidal ideation in adults with MDD [30, 31]. Preliminary research involving TMS to treat adolescents’ suicidal ideation has shown promising results [14]. Treatment with TMS most likely addresses pathological imbalances in prefrontal cortical GABAergic inhibitory function that underlie depression and suicidal ideation in adolescents [14,22,23,29,32]. Standard 10-Hz TMS, however, does not typically produce rapid relief from suicidal ideation and depressive symptoms [31,33,34].

A newer type of TMS, termed *theta burst stimulation* (TBS), induces changes in synaptic plasticity more rapidly than standard TMS [35–40]. Protocols with TBS appear to be safe in adolescents, are more tolerable than standard TMS, and are delivered over approximately 12-minute sessions vs 37-minute sessions with standard TMS [41, 42]. TBS was developed by mirroring endogenous hippocampal discharges [37]. In TBS dosing, 3-pulse, 50-Hz bursts are delivered every 200 ms (at 5 Hz). Stimulation with intermittent TBS has been shown to produce long-term potentiation-like effects [37,43,44], and continuous TBS reduces cortical excitability [37, 45]. In prior studies of sequential bilateral TBS, continuous TBS was delivered during each session to the right prefrontal cortex, followed by intermittent TBS delivered to the left prefrontal cortex [41, 46]. This technique is thought to target hypoactivity of the left DPFC and hyperactivity of the right DPFC associated with MDD and suicidal ideation [19,47,48]. Prior studies suggest that sequential bilateral TBS approaches clinically reduce depressive symptoms and suicidal ideation more than unilateral TBS [30,31,46].

Accelerated TBS (aTBS) dosing delivers multiple daily sessions of TBS [49]. The rationale for accelerated dosing is that multiple stimulations each day will produce faster, greater, and more durable effectiveness than standard, daily TMS sessions. Studies of aTBS dosing for adult patients with refractory depression have had encouraging results [36, 40]. Few prior studies have investigated the use of TBS and aTBS protocols in adolescents. However, the safety profile for adolescents appears to be similar to that for adults [41]. Prior study findings suggest that repeated TBS sessions safely enhance neurophysiological and clinical changes [41,42,50]. Recent work suggests that aTBS protocols more rapidly and effectively improve depressive symptoms and improve synaptic plasticity in the prefrontal cortex [36,39,40]. These aTBS protocols also overcome many practical limitations of standard TMS [49].

Integrating TMS with electromyography (EMG) or electroencephalography (EEG) in neurophysiological studies facilitates the study of cortical inhibition biomarkers and related neurotransmission [51–53]. Short-interval intracortical inhibition indexes cortical GABA_A_-mediated inhibitory neurotransmission, and long-interval intracortical inhibition (LICI) indexes the cortical GABA_B_-mediated inhibitory neurotransmission [53–55]. Data collected with TMS-EEG may yield more direct measures of prefrontal cortical function, but some contemporary methods are controversial [56–60], and TMS-EEG measures have not been reliably collected in adolescents with depression and suicidal ideation [59]. Conversely, with TMS-EMG, measures of prefrontal cortical function have been extensively studied in adolescents with depression and could easily be implemented in clinical environments [24,51,61–63]. Adolescents with MDD and suicidal behaviors have deficient LICI with related GABA_B_ impairment [25]. A preliminary study found that in adolescents treated for depression, LICI markers improve, with concurrent decrements in suicidal ideation [64]. Hence, TMS-EMG measures of LICI provide a promising biomarker for assessing and treating adolescents with depression and suicidal ideation.

### Objectives

The primary aim of this randomized controlled clinical trial is to compare the efficacy of sequential bilateral aTBS vs sham stimulation for treating suicidal ideation in adolescents with MDD. It is hypothesized that, compared with sham stimulation, sequential bilateral aTBS will provide greater improvements in intensity of suicidal ideation over the 10-day treatment course (hypothesis 1a) and over the 12-month follow-up period (hypothesis 1b) and would lead to fewer emergency department visits and fewer hospitalizations related to suicidality during 12-month follow-up (hypothesis 1c).

The secondary aim examines LICI, measured with TMS-EMG, as a predictive and target engagement biomarker for suicidality in adolescents with MDD. It is hypothesized cortical inhibition will demonstrate an indirect correlation with severity of suicidal ideation at baseline (hypothesis 2a) and over the 10-day treatment (hypothesis 2b). An exploratory aim examines the use TMS integrated with EEG (rather than EMG) to develop measures of cortical inhibition for assessing suicide risk and responsiveness to aTBS in adolescents with MDD (hypothesis 3). This concurrent TMS- EEG research will provide opportunities to further our understanding of the neurophysiological and molecular mechanisms of suicidal ideation in adolescents while refining biomarker methodology for future clinical use [60, 61].

## Methods

### Design and Study Setting

This is a randomized, double-blind, sham-controlled trial of sequential bilateral aTBS in adolescents with depression and suicidal ideation. This study follows an experimental medicine approach [65] implemented in an intensive outpatient program setting that supports local emergency departments, an 18-bed adolescent inpatient psychiatry unit, and outpatient clinics. The study is taking place at Mayo Clinic, Rochester, Minnesota, and the schedule of events is shown in the Figure.

#### Eligibility Criteria

This study is enrolling adolescents with MDD and suicidal ideation in both the treatment group and control (sham) group. Both inpatients and outpatients are eligible to participate if they meet the inclusion criteria shown in the Box and have none of the exclusion criteria listed there. Use of antidepressant medication (if recommended by the referring clinician and agreed on by the parent/guardian and patient) is allowed.

#### Intervention

The TMS system (MagVenture TMS Therapy, MagVenture Inc) used in this study is cleared by the US Food and Drug Administration (FDA) for treatment of MDD in adult patients who did not receive satisfactory improvement from antidepressant medication in the current depressive episode. This TMS protocol delivers intermittent TBS for 3 minutes and is based on a patterned TMS paradigm delivered to the left DPFC at 120% resting motor threshold (MT). It consists of bursts of 3 pulses at 50 Hz with bursts repeated at 5 Hz in a duty cycle of 2 seconds on, 8 seconds off over 3 minutes 9 seconds, for a total of 600 pulses. Treatment typically is administered daily for 5 days per week, for 4 to 6 weeks.

In the present study, TBS will be delivered on both the right and left DPFC, 3 times daily for 10 days (5 days per week). During each session, *continuous* TBS, in which 1,800 pulses will be delivered continuously over 120 seconds, is administered first to the right DPFC. Then intermittent TBS is administered to the left DPFC, with 1,800 pulses delivered in 2-second bursts and repeated every 10 seconds for 570 seconds. The comparison group will receive 3 daily sessions of bilateral sham TBS for 10 days. The system has both active and placebo (sham) coils (MagVenture Cool-B70 A/P coils, MagVenture Inc). Because of the nature of the accelerated treatment paradigm and the age of the target population (13-18 years), the intensity will be decreased from the usual 120% MT to 80% MT. The TBS parameters were adopted from prior research [41].

During the trial, all participants will receive standard-of-care treatment of depression and suicidal ideation, including daily psychotherapeutic skill sessions during TBS therapy [7, 66]. The 10-day psychotherapy treatment program was adapted from a youth suicide prevention program [66].

#### Discontinuing or Modifying Allocated Interventions

Participants may withdraw voluntarily from the study at any time. The participant stopping rules apply during the 2-week period when the participants are receiving the study intervention (active or sham sequential bilateral aTBS). These rules do not apply to participants in the naturalistic 1- year follow-up portion of the study. Participants will be withdrawn from the study by the PI (Principal Investigator) if they have any of the following:

- A seizure
- Treatment-induced mania as confirmed by *Diagnostic and Statistical Manual of Mental Disorders* (Fifth Edition) definitions
- Psychotic symptoms
- Noncompliance with study procedures
- Illicit drug use or a positive urine drug screen during the study

#### Strategies to Improve Adherence to Interventions

All participants will be provided with an intensive, 10-day psychotherapeutic intervention during the trial of sequential bilateral aTBS [41,66,67]. This psychotherapeutic program meets and exceeds both local and national standards of care for depressed adolescents with suicidal ideation.

Our prior experience suggests that the sequential bilateral aTBS protocol in the present study will facilitate participant retention because it is a more pragmatic schedule with 10 treatment days total. Mayo Clinic’s neuromodulation research team can offer treatment sessions over the weekend if necessary to accommodate an adolescent’s or family’s schedule.

#### Concomitant Care During the Trial

During the trial, participants will continue to take antidepressant medication if recommended by their referring clinician. Participants are not required to take antidepressant medication for study participation because of ethical, human subject protection, and practical concerns. During the treatment phase, zaleplon, zolpidem, or zopiclone (1 dose nightly) as needed for treatment-emergent insomnia or lorazepam (up to 2 mg/d) for treatment- emergent anxiety may be administered for up to 10 doses. The use of alternative hypnotics or anxiolytic compounds requires prior approval from the PI. Hormonal contraceptives are allowed. Short-term treatments of headache, allergy, viral rhinitis, and influenza symptoms will be allowed during the trial. All questions regarding the acceptability of specific medications must be answered by the PI.

#### Provisions for Posttrial Care After Withdrawal

The PI will assist with clinical referrals for all participants who voluntarily withdraw from the study or are withdrawn by the research team and PI. Specifically, the PI will offer ongoing clinical care or community referrals depending on the participant’s preference.

### Outcomes

The primary outcome is efficacy of sequential bilateral aTBS vs sham stimulation for reducing suicidal ideation in adolescents with MDD. The primary acute outcome measure is change in the score of the Columbia– Suicide Severity Rating Scale (C-SSRS) severity of ideation subscale [68] over the 10-day treatment. Additional outcome measures include monthly follow-up C-SSRS severity of ideation subscale scores, number of hospitalizations related to suicidal ideation, and number of emergency department visits related to suicidal ideation over the 12-month follow-up period.

The secondary outcome is to examine LICI as a predictive and target-engagement biomarker for suicidality in adolescents with MDD. Suicidal ideation will be measured with the C-SSRS severity of ideation subscale, and cortical inhibition will be assessed by using LICI at an interstimulus interval of 100 ms (LICI-100) collected with TMS-EMG at baseline and at the end of the 10-day treatment.

An exploratory outcome is developing TMS-EEG measures of cortical inhibition (TMS-EEG at 45 ms after stimulation [N45] and 100 ms after stimulation [N100]) for assessing suicide risk and responsiveness to aTBS in adolescents with MDD.

### Participant Timeline

The schedule of enrollment, interventions, assessments, and visits is shown in Table 1. The first visit for study participants is a screening visit. At this visit, informed consent and assent will be completed. A medical history will be taken, vital signs (pulse, blood pressure, temperature, height, and weight) will be measured, and a physical examination, urine pregnancy test (if applicable), and urine drug screen will be completed. The participant’s parent or guardian will not be told the pregnancy test result without the participant’s permission, but if the study physician believes that the pregnancy may cause serious health problems, they may need to tell the parent or guardian the test results. Patients with a positive urine drug screen that is not due to prescribed medications are ineligible for study participation. The study physician will ask if the results can be shared with the parent or guardian. This information will be kept confidential if the participant chooses not to share the drug test results with the parent or guardian. The result from the urine drug test will not become part of the medical record, but the results will remain in the study record.

**Table 1.**
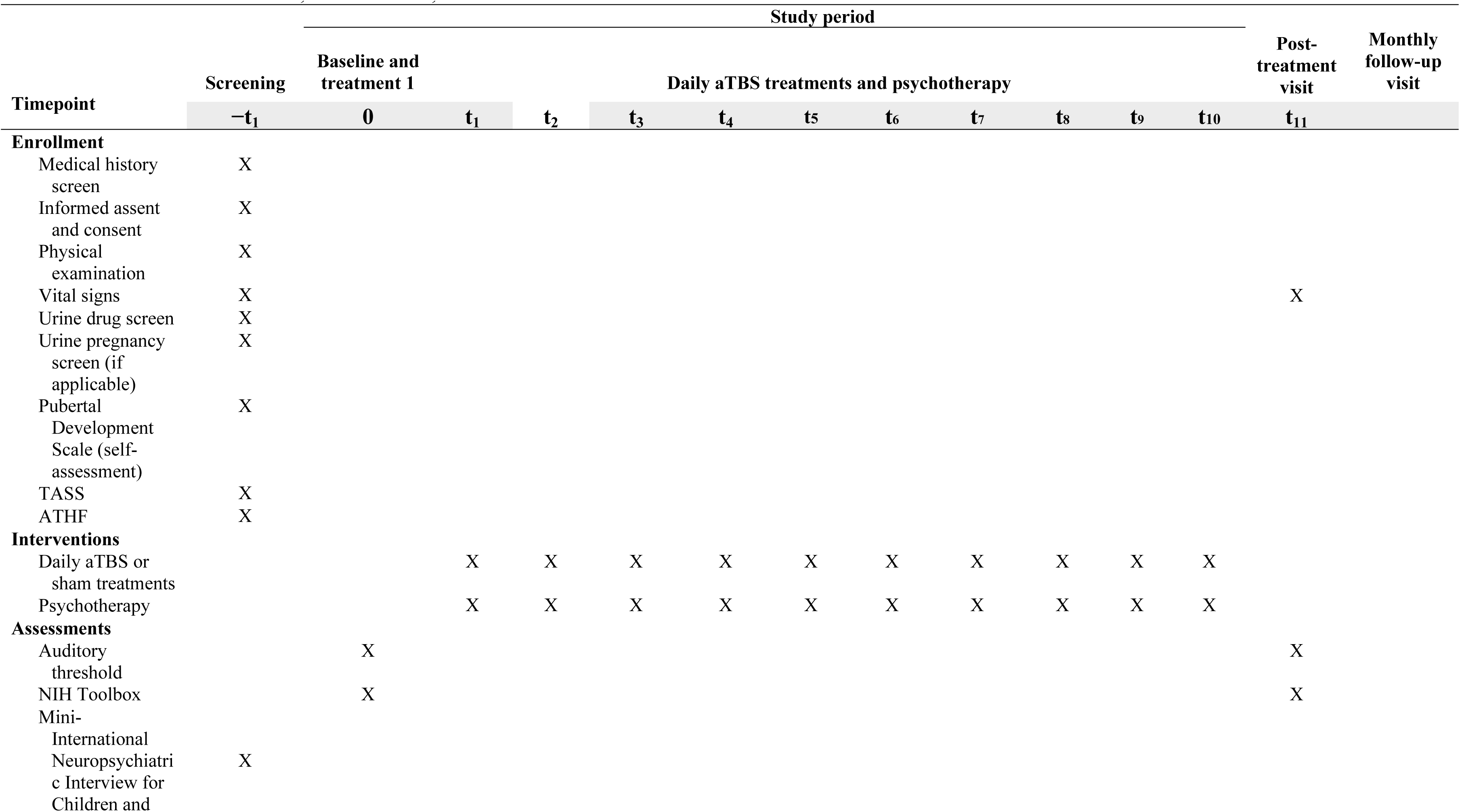
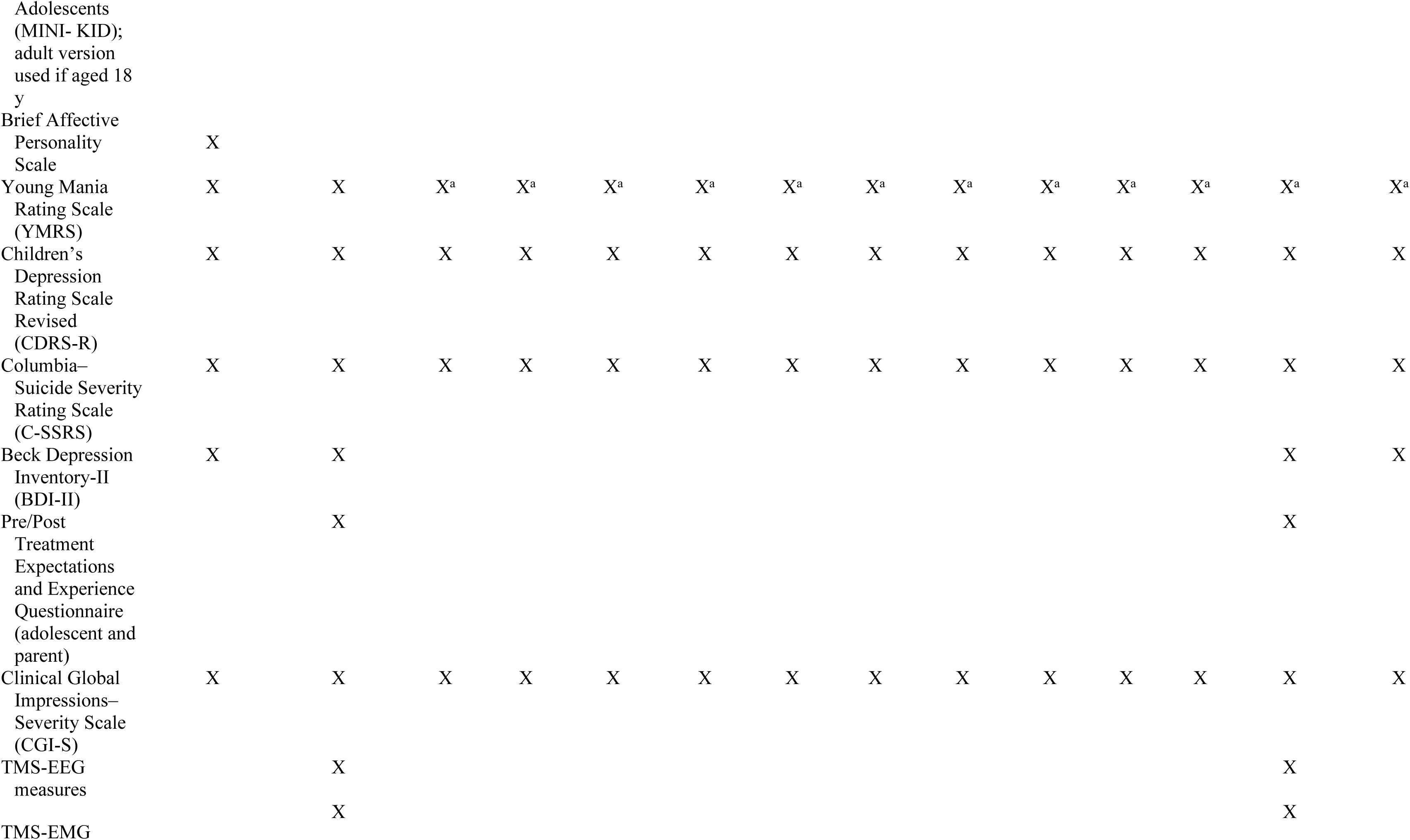
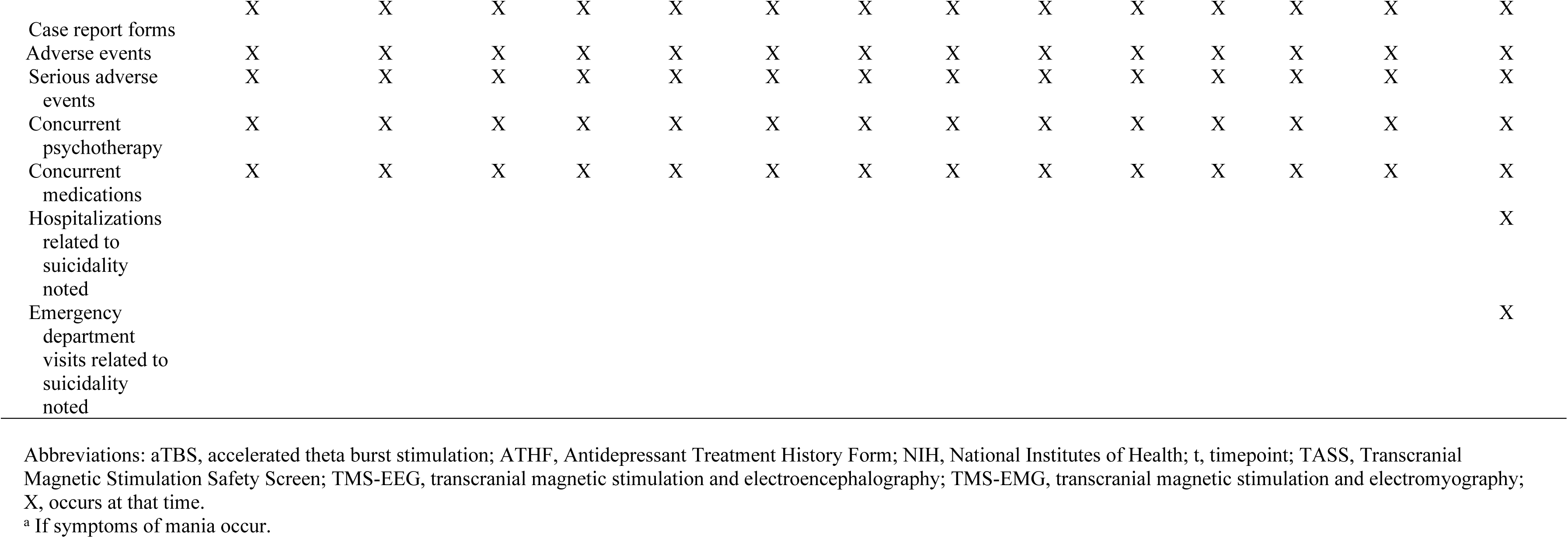
Schedule of Enrollment, Interventions, and Assessments

Additional screening assessments (Table 2) include the Transcranial Magnetic Stimulation Safety Screen [69], Antidepressant Treatment History Form [70], Pubertal Development Scale [71], Mini- International Neuropsychiatric Interview for Children and Adolescents (MINI-KID) for participants aged 12 to 17 years [72] or the Mini- International Neuropsychiatric Interview for participants aged 18 years [73], Young Mania Rating Scale [74], Children’s Depression Rating Scale Revised (CDRS-R) [75], C-SSRS [68], Beck Depression Interview-II [76], and Clinical Global Impressions–Severity Scale [77]. Also discussed and noted at the screening visit are concurrent use of medications, adverse events (AEs), serious AEs (SAEs), the case report form (CRF), and concurrent psychotherapy. The screening visit can occur on the same day or across 2 consecutive days depending on participant preference.

**Table 2.** Description of Assessments

| **Assessment** | **Description** |
| --- | --- |
| Antidepressant Treatment History Form (ATHF) | Systematic documentation of prior psychotropic medication exposure and to quantify treatment resistance (No. of failed antidepressant trials with an adequate dose and duration) |
| Auditory thresholds | Audiometry testing |
| Beck Depression Inventory-II (BDI-II) | Validated, 21-item, self-report measure with a score range of 0-63 |
| Brief Affective Neuroscience Personality Scale | Validated 33-item, self-report measure collected to assess behavioral characteristics in 6 affective neurobiological systems (play, seek, care, fear, anger, sadness) |
| Children's Depression Rating Scale Revised (CDRS-R) | Validated, 17-item, clinician-rated tool to assess severity of depression, with total possible scores ranging from 17 to 113; parents provide input into 14 of the items |
| Clinical Global Impressions– Severity Scale (CGI-S) | A standard measure of symptom severity and treatment response |
| Columbia–Suicide Severity Rating Scale (C-SSRS) | Validated, clinician-rated tool collected to assess lifetime and ongoing suicidal ideation and behavior; scale's severity of ideation subscale is a 5-point ordinal scale that is the primary clinical outcome measure for this study |
| Mini-International Neuropsychiatric Interview for Children and Adolescents (MINI-KID) and Mini-International Neuropsychiatric Interview (MINI) | Structured psychiatric interview (MINI-KID for ages 12-17 y and MINI for $\geq 18$ y) done to confirm diagnostic inclusion criteria and to rule out diagnostic exclusion criteria |
| NIH Toolbox Cognition Battery | Battery of cognitive tests recommended for ages $\geq 7$ y; consists of tests of multiple constructs; it yields individual measure scores and these summary scores: Education, Handedness, Attention (Flanker Inhibitory Control and Attention Test), Episodic Memory (Picture Sequence Memory Test, and Auditory Verbal Learning Test), Executive Function (Dimensional Change Card Sort Test, Flanker Inhibitory Control and Attention Test), Language (Picture Vocabulary Test, Oral Reading Recognition Test), Processing Speed (Pattern Comparison Processing Speed Test), Working Memory (List Sorting Working Memory Test) |
| Pre/Post Treatment Expectations and Experience Questionnaire (adolescent and parent) | Assessment of patient and parent impressions and expectations and integrity of blinding by asking adolescents and parents to guess their study arm assignment |
| Pubertal Development Scale and Tanner staging | Self-report questionnaire and line drawings used to evaluate level of pubertal development |
| Transcranial Magnetic Stimulation Safety Screen (TASS) | Safety screen completed before TMS to confirm patient safety for TMS |
| Young Mania Rating Scale (YMRS) | An 11-item measure used to assess for manic symptoms; if the YMRS is positive, a MINI or MINI-KID will be repeated to evaluate for mania, and if the study participant meets full *DSM-5* criteria for mania or hypomania, that person will be discontinued from the study and followed up clinically |
| TMS neurophysiology measures | TMS-EMG (LICI-100) and TMS-EEG (N45 and N100) neurophysiology measures (biomarkers of cortical inhibition and activity) |

Abbreviations: aTBS, accelerated theta burst stimulation; *DSM-5*, *Diagnostic and* *Statistical Manual of Mental Disorders* (Fifth Edition); LICI-100, long-interval intracortical inhibition at an interstimulus interval of 100 ms; NIH, National Institutes of Health; N45, 45 ms after stimulation; N100, 100 ms after stimulation; TMS, transcranial magnetic stimulation; TMS-EEG, TMS and electroencephalography; TMS-EMG, TMS and electromyography.

After eligibility is confirmed, additional assessments will be completed at a subsequent visit. These assessments include the NIH (US National Institutes of Health) Toolbox Cognition Battery [78] and a pretreatment expectations survey. The Young Mania Rating Scale, CDRS-R, C-SSRS, and Beck Depression Interview-II will be repeated, and concurrent use of medications, concurrent psychotherapy, and CRFs will be reviewed. During the intervention period, AEs and SAEs will be recorded. TMS neurophysiological data also will be collected.

After acceptance into the trial, each participant is randomly assigned to either the active aTBS group or sham aTBS, as described in the Randomization and Blinding section. Coil localization (Beam F3 method) and MT procedures are completed [79]. The participant will complete day 1 of aTBS as assigned (either 3 active sequential bilateral TBS sessions or 3 bilateral sham sessions). Treatment continues for 9 more days if tolerated. ***Sample Size***

Based on prior experience, it is anticipated that 200 patients will be screened and 100 will be enrolled to reach the target sample size of 80 needed to complete this pilot study. The sample size for the primary outcome (suicidal ideation over 10 days of sequential bilateral aTBS treatment) was estimated by incorporating a repeated-measures design with assumptions about potential participants (eg, effect size, odds ratio [OR]). These assumptions were based on a 2018 study of the effect of bilateral repetitive TMS vs sham stimulation on suicidal ideation in adult patients with treatment-resistant MDD [31]. The results from the a priori sample size estimation and power analysis suggested that a sample size of 35 participants per group (N=70) would achieve 80% power at a .05 α level (2-tailed) [31]. The aim is to detect an overall OR of 2.85 for the primary outcome of less suicidal ideation with sequential bilateral aTBS vs sham stimulation in a 2- group (between-participants) randomized design. The posited OR is based on 10 (within-participants) repeated measurements of severity of suicidal ideation on the CSSR-S subscale, when the proportion (incident rate) of suicidal ideation from the sham group at the end of the 10-day trial meets or exceeds 0.50 and the assumed within-participant correlation between observations (suicide ideation) for the same participant is 0.50 or less. (A correlation of 0.50 was based on guidance from our preliminary study of TMS [14].) The posited OR of 2.85 for the current application is slightly more conservative than what was reported in a study by Weissman et al [31] of effects of bilateral TMS vs sham stimulation on suicidal ideation in adult patients with treatment-resistant MDD (OR=3.03).

The dropout rate is estimated to be 10%, an approximation based on treatment discontinuation results from our previous repetitive TMS studies [14,80,81]. Thus, 80 participants will be enrolled to allow for this expected rate of attrition with the intent of capturing evaluable data for 70 participants. PASS (Power Analysis and Sample Size) 2019 software version 14.0.15 (NCSS) was used to perform the sample size and power analysis.

To definitively evaluate the effect of sequential bilateral aTBS treatment on suicidal ideation in adolescents, we may require a sample size larger than 70. The treatment effect of sequential bilateral aTBS has not been previously studied in adolescents with suicidal depression, and we have no previous findings from an adolescent population to guide effect size selection for the present effort. The selection of 70 participants was, in part, resource driven. The sample size is designed to detect a moderate effect and to establish a proof of concept so that the research can expand to a subsequent, larger-scale multicenter study. Nevertheless, the current pilot study permits an evaluation of whether receiving sequential bilateral aTBS shows initial evidence of improving suicidal ideation in depressed adolescents.

### Recruitment

Recruitment began April 4, 2022, after the protocol v2.0 was finalized and approved by the Data and Safety Monitoring Board (DSMB) November 10, 2021. The estimated completion date of the study is December 31, 2026. Adolescent participants are being recruited through the Mayo Clinic Depression Center and Division of Child and Adolescent Psychiatry in Rochester, Minnesota. Recruitment is occurring through outpatient clinics, an 18-bed child and adolescent inpatient psychiatry unit, and Mayo Clinic Emergency Medicine Department in Rochester. Mayo Clinic Health System clinics throughout Minnesota and Wisconsin provide additional recruitment environments. Ongoing and future outreach efforts also involve local community and private treatment facilities in Rochester, St Paul, and Minneapolis in Minnesota. The PI (P.E.C.) has prior and planned collaborations with the Rochester Public School District, Rochester, Minnesota, including consultation with the school board. With appropriate approvals, recruitment flyers will be disseminated to the parents of all adolescents attending Rochester public schools.

During inpatient hospitalizations, outpatient visits, and emergency medicine department evaluations, a treatment team member will inform the research team and attending physician of a patient’s potential interest. The attending physician will review the request and determine whether a discussion of research participation is appropriate. The PI or another research team member will be contacted to speak to the patient and family regarding participation. If the patient and family are interested, the research team will initiate the informed consent process.

The research team will discuss the study during staff meetings at internal clinical services and external facilities. External efforts will be tailored to foster community education regarding depression and suicidality in adolescents while establishing referral patterns. Flyers approved by the Mayo Clinic Institutional Review Board (IRB) will be posted in facilities to advertise the study. The research team will participate in local and national events such as Suicide Awareness Day, National Depression Day, and National Mental Health Day to facilitate community engagement and participant recruitment. The PI will continue to access and collaborate with relevant offices in the Mayo Clinic Center for Clinical and Translational Science such as the Office of Community Engagement. These collaborations provide local speaking opportunities.

### Randomization and Blinding

Randomization will occur for each participant when that person meets all inclusion criteria, does not have any exclusions, and has satisfactorily completed all prestudy procedures. The study biostatistician (P.A.N.) generated a randomization schedule using a random number program on software (SAS via PROC PLAN, SAS Institute Inc). Participants will be randomly assigned in a 1:1 ratio to receive 1 of the 2 treatment regimens (either 3 daily sequential bilateral aTBS sessions in the active arm or 3 daily sham sessions in the control arm) for 10 days.

The use of active and placebo (sham) coils facilitates blinding of the operator and participants receiving aTBS or sham treatments. A pretreament and posttreatment expectancy and experience form will ask participants and parents to make a guess regarding their treatment arm (active or sham). Blinded study team members will also be asked to guess treatment assignments for each participant. The study team members completing clinical rating scales will not be allowed in the treatment suite during TBS treatment sessions and will be blinded to treatment assignment.

In the event of a device-related serious adverse effect, the PI will carefully assess whether breaking the blinding will critically affect how a participant is treated in response to the adverse effect and whether this knowledge outweighs the implications to the scientific soundness of the study. This decision will be reviewed and approved by the study medical monitor. For most serious adverse effects, treatment would be discontinued, and symptoms would be treated regardless of the knowledge of whether the treatment received was active or sham.

### Data Collection and Management

The primary acute outcome measure is the score on the C-SSRS of the 5-point severity of ideation subscale, with 1 indicating wish to be dead; 2, nonspecific active suicidal thoughts; 3, suicidal thoughts with any methods (no plan) without intent; 4, active suicidal ideation with some suicidal intent without a plan; and 5, active suicidal ideation with suicidal intent and a plan [68]. Another C-SSRS subscale that will be used is the intensity of ideation subscale (composed of frequency, duration, controllability, deterrents, and reasons for ideation) [68]. The C-SSRS has been validated for assessing suicidal ideation and behavior in adolescents and adults, with good predictive validity for death by suicide [68] (Table 2). The other study instruments are mentioned in the Participant Timeline section and Table 2.

Study retention is expected to be high for this study for multiple reasons. During the treatment phase of the study, participants will have frequent contact with the study team’s child and adolescent psychiatrist (10 daily visits). Both the participants and their parent or legal guardian will be told to contact their study doctor if the participant has any major mood or behavioral changes including suicidal ideation. Ongoing monitoring for worsening of depression and emergence of suicidal ideation and suicidal behaviors will be performed with the CDRS-R, C-SSRS, and Clinical Global Impression-Severity Scale [68,77,82]. Additionally, the scheduling flexibility (including weekends) and the TBS dosing parameters (more tolerable treatment intensity) should be highly effective in retaining adolescent participants in this study. Nonadherence to the assigned treatment regimen is defined as missing 2 or more treatment days, or missing more than 20% of the total number of treatment sessions (ie, 6 sessions) occurring during the 2 weeks of treatment as outlined in the schedule of events. Outcomes data will be included in the analyses for any participant randomly assigned into the study who received at least 1 exposure (treatment) from the study device.

A CRF will be completed for each participant enrolled in the clinical study. The investigator-sponsor will review, approve, and sign and date each completed form to show responsibility for ensuring that all clinical data entered on the CRF are complete, accurate, and authentic. All study data will be managed as follows:

1. Data will be entered into a secure database (REDCap, Vanderbilt University).
2. Study data will be reviewed regularly by the PI and the Study Coordinator for the following:

a. Inclusion criteria are met.
b. Data are transcribed accurately and completely.
c. Units of measure are recorded appropriately.
3. Details of the study treatments including treatment parameters, such as percentage MT for each treatment, will be stored in the device manufacturer’s software (MagVenture Research Study System software, MagVenture Inc). A daily summary of the treatments will be created and stored in the participant’s source document.
4. Case files will be created for each participant where completed visit CRFs will be stored. Any AE-related CRFs will also be stored in the case file.
5. Nonelectronic source documents containing data will be stored in a locked cabinet in a secure office. Only authorized study staff will have access.
6. Electronic data will be stored in a secure database. Only authorized users will have access. All users will have unique identifiers and passwords. Sharing logon information is not permitted.

### Statistical Analysis

All statistical analyses will be performed with SAS software version 9.4 (SAS Institute Inc). The level of significance will be set at α=.05 (2-tailed). To address multiple testing where applicable, *P* values will be adjusted by using the false discovery rate [83].

Suicidal ideation (as measured by the score of the C-SSRS severity of ideation subscale) over the 10-day sequential bilateral aTBS trial period is the primary ordinal outcome measure (hypothesis 1a). The short- term change over time (10 days) in suicidal ideation will be compared between the 2 treatment groups (sequential bilateral aTBS vs sham stimulation) by using an ordinal logistic regression model within a generalized estimating equation (GEE) framework. The logistic model will contain fixed-effects terms for treatment, time, and treatment × time interaction along with baseline C-SSRS severity of ideation subscale as a covariate. Age, sex, pubertal status, antidepressant medication status, and depression severity (CDRS-R total as a time-varying covariate) will also be covariates in the model. Simple treatment group effects will also be assessed in each study period (10 days or 12 months). (See the sensitivity analysis section later for a missing not at random [MNAR] mechanism.)

After the treatment period ends, suicidal ideation will be assessed at monthly follow-up visits over a 12-month period (hypothesis 1b). An ordinal logistic regression model within a GEE analysis framework similar to that described for the primary outcome (hypothesis 1a) will be used to estimate suicidal ideation over the 12-month follow-up period. In addition to the covariates specified for the primary outcome, we will also consider including pharmacotherapy and psychotherapeutic intervention received over the follow-up period as additional covariates.

Emergency department visits and hospitalization visits related to suicidality will be assessed at monthly follow-up visits over the 12-month follow-up period (hypothesis 1c). A separate Poisson mixed-model analysis will be used to estimate the number of emergency department visits and the number of hospitalization visits for those who received sequential bilateral aTBS and for those who received sham stimulation. The model will contain fixed-effects terms for treatment, time, and treatment × time interaction, with the same covariates identified earlier. Simple treatment group effects will also be assessed in each period. Maximum likelihood estimation, type 3 tests of fixed effects, and generalized least-squares means will be used along with the best-fitting covariance structure. The sandwich (robust covariance matrix) estimator will also be considered. The Poisson model will be checked for overdispersion. The Poisson regression will model the logarithm of each outcome as a random variable, and the mean and variance will be estimated on the logarithmic scale. Thus, to facilitate interpretation, geometric means will be reported, which will be obtained by exponentiating the log-scale parameter estimates. Generalized linear mixed-model procedures software (PROC GLIMMIX, SAS/STAT, SAS Institute Inc) will be used to conduct the Poisson regression for hypothesis 1c and provide a robust mechanism for handling data that are assumed missing at random. (See the sensitivity analysis section for data that are MNAR.) No subgroup analyses are planned.

An ordinal logistic regression model will be used to estimate severity of suicidal ideation at baseline from LICI-100 measured with TMS- EMG before the intervention (hypothesis 2a). Age, sex, pubertal status, antidepressant medication status, and depression severity at baseline (CDRS- R total) will be covariates in the model. The maximum likelihood estimation and type 3 tests of fixed effects will be used along with the Wald χ^2^ statistic. Robust standard errors (Huber sandwich estimator) will also be considered. For the ordinal logistic regression, the cumulative probabilities will be modeled over the lower-ordered suicidal ideation scale scores (less suicidal ideation). The adjusted ORs (odds of suicidal ideation from LICI with TMS- EMG) and 95% CI will be estimated as part of the model to interpret the relationship between the TMS-EMG biomarker and baseline suicidal ideation.

An ordinal logistic regression model within a GEE analysis framework similar to that described for hypothesis 1a will be used to examine the relationship between cortical inhibition (LICI-100 measured with TMS-EMG as a time-varying covariate) and C-SSRS severity of ideation subscale (as the response variable) over the 10-day treatment trial (hypothesis 2b). The parameter estimates (regression coefficients) will be interpreted from the solution for fixed effects in the GEE analysis. The adjusted OR (odds of suicidal ideation from cortical inhibition) and 95% CI will be estimated as part of the GEE model. The same covariates specified for hypothesis 1a will be in the model for hypothesis 2b.

An ordinal logistic regression model within a GEE analysis framework similar to that described earlier for hypothesis 2b will be used to examine the relationship between cortical inhibition (TMS-EEG markers N45 and N100) and C-SSRS severity of ideation subscale (as the response variable) over the 10-day treatment trial (hypothesis 3). A separate model will be implemented for each measure of TMS-EEG cortical inhibition. The same covariates specified for hypothesis 2b will be in the model for hypothesis 3. The parameter estimates (regression coefficients) will be interpreted from the solution for fixed effects in the GEE analysis. The adjusted OR (odds of suicidal ideation from TMS-EEG cortical inhibition) and 95% CI will be estimated as part of the GEE model.

#### Sensitivity Analysis to Handle Missing Data

A sensitivity analysis is planned to address incomplete response patterns according to an MNAR mechanism. First, we will follow the outcome-based model of Diggle and Kenward [84] for longitudinal data with nonrandom missingness along with the model-based approach proposed by Carpenter et al [85] for modeling both the treatment response and the missing data process. These models are summarized here. If deemed necessary, we will build longitudinal models for the data, which include a model for the dropout (or missing data) process to assess the sensitivity of the results about the dependence between dropout (or missingness) and treatment response. Additionally, if deemed necessary, we will consider implementing the semiparametric shared parameter model described by Tsonaka et al [86] to handle nonmonotone nonignorable missingness for mixed-effects models. A pattern-mixture model will also be considered if necessary. The results from such nonrandom dropout/missingness models will then be compared with the results from the proposed standard methods (dropout or missing at random models) to assess the sensitivity of the results to the effect of dropouts (or incomplete response patterns according to an MNAR mechanism).

### Data Monitoring

An National Institute of Mental Health (NIMH)-constituted DSMB is overseeing the study in accordance with the NIMH DSMB charter guidelines [87]. Clinical monitoring will be conducted by the NIMH Clinical Research Education, Support, and Training program. Goals of clinical site monitoring are to ensure that the rights and well-being of participants are protected; that the reported trial data are accurate, complete, and verifiable; and that the conduct of the trial complies with the approved protocol and any amendments, good clinical practice, and applicable regulatory requirements. Clinical monitoring includes reviewing the study database and documents including regulatory documents. Monitoring will be conducted throughout the study and involves targeted data verification of key data variables.

Details of clinical site monitoring are documented in the clinical monitoring plan. This plan describes who will conduct the monitoring, at what frequency monitoring will be done, at what level of detail monitoring will be performed, and the distribution of monitoring reports. Copies of monitoring reports will be provided to the PI and to the NIMH DSMB liaison.

An individualized quality management plan was developed to describe the clinical site’s quality management in addition to the site’s assurance and quality control measures already in place. The site staff will perform internal quality management of study conduct, data collection, and data documentation.

#### Interim Analyses and Study-Stopping Rules

No interim analyses are planned. Study-stopping rules were developed in collaboration with the FDA and NIMH DSMB to ensure that participants are not exposed to risks across the study and that any potential risks are evaluated. These stopping rules apply to the 2-week period when participants are receiving the study intervention (active or sham sequential bilateral aTBS) but do not apply to the naturalistic 1-year follow-up portion of the study. No study-stopping rules were predetermined for events that take place during the follow-up period because suicide attempts and psychiatric hospitalizations are expected to be common events. AEs occurring during the follow-up period will be tracked. The study team will evaluate the expectedness and relatedness of these events; the IRB, DSMB, and FDA will provide oversight of these determinations. AEs and SAEs will be reported in the data reports and reviewed by the DSMB. The following study-stopping rules apply:

- Two participants who experience a seizure.
- One or more participants who experience status epilepticus.
- One or more participants who attempt or die by suicide.
- Two or more SAEs related to the study intervention and occurring within the 2-week treatment period for 1 or more participants.

If these study-stopping rules are met, enrollment of new participants will cease, but all study activities will continue for enrolled participants. The PI will consult with the DSMB and the FDA within 10 business days of initial notice that study-stopping rules were met.

#### AE Reporting and Harm

All AEs occurring during the study, including those not meeting the criteria of an unanticipated adverse device effect (UADE), will be recorded on the appropriate CRF. Expected clinical AEs and anticipated adverse device effects include pain or discomfort under the treatment coil; eye, facial, or skin pain; facial numbness or facial muscle twitching; toothache; blurred vision; seizures; cardiogenic syncope; sedation; and worsening mood symptoms. UADEs will be reported to the DSMB, FDA, and IRB immediately. Records of these events will be maintained, and reports will be submitted to the FDA and IRB according to the regulatory requirements. Expected clinical AEs and nonserious clinical AEs will be reported annually.

The study follow-up period is defined as 12 months after the last administration of study treatment. All AEs occurring during the study treatment period will be recorded as required. All observed or volunteered adverse effects (serious or nonserious) and abnormal test findings will be recorded in the participant’s case history regardless of the treatment group or a suspected causal relationship to the investigational device or, if applicable, to other study treatment or diagnostic products. For all adverse effects, sufficient information will be pursued and obtained regarding permit, an adequate determination of the outcome, and an assessment of the causal relationship between the adverse effect and the investigational device or other study treatment or diagnostic product. The clinical course of each event will be followed up, as required, until resolution or stabilization or until the study treatment or participation is determined to not be the probable cause. SAEs that are still ongoing at the end of the study will be followed up to determine the final outcome. Any SAE that occurs after the study period and is considered to be at least possibly related to the study treatment or study participation will be recorded and reported immediately.

The sponsor-investigator will promptly review documented adverse effects and abnormal test findings to determine the following: 1) whether the abnormal test finding should be classified as an adverse effect; 2) whether there is a reasonable possibility that the adverse effect was caused by the investigational device or other study treatments; and 3) whether the adverse effect meets the criteria for a serious adverse effect. If the sponsor-investigator’s final determination of causality is “unknown and of questionable relationship to the investigational device or other study treatments,” the adverse effect will be classified as associated with the use of the investigational device or other study treatments for reporting purposes. If the sponsor-investigator’s final determination of causality is “unknown but not related to the investigational device or other study treatments,” this determination and the rationale for the determination will be documented in the respective participant’s case history.

The sponsor-investigator will promptly review documented UADEs (FDA Form 3500A) and will report the results from the evaluation to the FDA and DSMB within 10 workdays and to the Mayo Clinic IRB within 5 workdays of initial notice of the effect. Thereafter, the sponsor- investigator will submit additional reports concerning the effect as requested.

The sponsor-investigator will permit study-related monitoring, audits, and inspections by the IRB, the monitors, and government regulatory agencies of all study-related documents (eg, source documents, regulatory documents, data collection instruments, study data). The sponsor- investigator will ensure the capability for inspections of applicable study- related facilities (eg, pharmacy, diagnostic laboratory). Participation as a sponsor-investigator in this study implies acceptance of potential inspection by government regulatory authorities and applicable compliance offices. ***Ethics and Dissemination***

#### Informed Consent

Prospective participants will be assured that participation is voluntary. All participants for this study will receive a consent/assent form describing this study and providing sufficient information for participants and their parents or legal guardian to make an informed decision about whether to participate. Formal consent must be obtained by using the approved IRB consent form before a participant undergoes any study procedure. The consent form must be signed and dated by the participant or the participant’s legally authorized representative, and by the study team member obtaining the informed consent.

#### Confidentiality

Information including clinical information about study participants will be kept confidential and managed according to the requirements of the Health Insurance Portability and Accountability Act of 1996 (HIPAA). Those regulations require a signed authorization by each study participant informing the participant of the following: what protected health information (PHI) [12] will be collected from participants, who will have access to that information and why, who will use or disclose that information, and the rights of a participant to revoke their authorization to use their PHI. If a participant revokes authorization to collect or use PHI, the investigator, by regulation, retains the ability to use all information collected before the revocation of participant authorization. For participants who have revoked authorization to collect or use PHI, attempts will be made to obtain permission to collect at least vital status (long-term survival status that the participant is alive) at the end of the originally scheduled study period.

Participant confidentiality and privacy are strictly held in trust by the participating investigators, their staff, and the sponsor. Therefore, the study documentation, data, and all other information generated will be held in strict confidence. No information concerning either the study or the data will be released to any unauthorized third party without prior written approval of the sponsor. All research activities will be conducted in as private a setting as possible. Potential participants will have the opportunity to discuss study participation without their parent or guardian present. The study monitor, other authorized representatives of the sponsor, and representatives of the NIH, the FDA, and the Mayo Clinic IRB may inspect all documents and records required to be maintained by the investigator, including but not limited to medical records. Study participants’ contact information will be securely stored for internal use during the study. At the end of the study, all records will continue to be kept in a secure location for as long as dictated by the reviewing IRB, institutional policies, FDA investigational device exemption (IDE) record requirements, and sponsor requirements. We are not planning to collect biological specimens.

To further protect the privacy of study participants, the NIH will issue a Certificate of Confidentiality. This certificate protects identifiable research information from forced disclosure. It allows the investigator and others who have access to research records to refuse to disclose identifying information on research participation in any civil, criminal, administrative, legislative, or other proceeding, whether at the federal, state, or local level. By protecting researchers and institutions from being compelled to disclose information that would identify research participants, Certificates of Confidentiality help achieve the research objectives and promote participation in studies by helping assure confidentiality and privacy to participants.

#### Dissemination

The PI/sponsor holds the primary responsibility for publications of results from the proposed study. The PI/sponsor and study team will present findings at national and international conferences and will publish findings in peer-reviewed journals. Results will be posted to ClinicalTrials.gov within 12 months of final data collection for the primary outcomes.

## Discussion

This is the first known randomized controlled trial to examine sequential bilateral aTBS in depressed adolescents with suicidal ideation. The study addresses 2 NIMH Strategic Research Strategies: 3.2, “Develop ways to tailor existing and new interventions to optimize outcomes,” and 3.3, “Test interventions for effectiveness in community practice settings” [88, 89]. If this aTBS treatment protocol and the proposed panel of biomarkers are shown to be safe and effective, they can be rapidly implemented into clinical practice. This novel approach may facilitate a critical paradigm shift toward brain-based approaches for assessing and treating suicidal ideation in adolescents with MDD.

## Data Availability

No datasets were generated or analysed during the current study. All relevant data from this study will be made available upon study completion.

## Supplementary Information

The online version contains supplementary material (Appendix 1. Standard Protocol Items: Recommendations for Interventional Trials [SPIRIT] 2013 Checklist [additional file 1.doc]; Appendix 2. Study Administrative Information [additional file 2.doc]. Appendix 3. Sample Consent Forms [additional file 3.pdf and additional file 4.pdf] [Used with permission of Mayo Foundation for Medical Education and Research.].

### Declarations

#### Ethics Approval and Consent to Participate

This study is to be conducted according to US government regulations and institutional research policies and procedures. This protocol was submitted, and any amendments will be submitted, to a properly constituted local institutional review board (IRB), the Mayo Clinic IRB, in agreement with local legal prescriptions, for formal approval of the study. The IRB approved this study (#20-009630). All participants or their parents or legal guardian will be asked to sign an informed consent form as described in the article.

#### Consent for Publication

Not applicable.

#### Availability of Data

Because the study is funded by the National Institute of Mental Health (NIMH), data sharing through the NIMH Data Archive (NDA) is required. The NDA is a large database in which deidentified study data from many NIMH studies are stored and managed. Any researcher who requests access to the deidentified data of the NDA is bound to adhere to strict data safety practices and to avoid any attempts at deidentification. The investigators’ intention to submit deidentified participant data to the NDA is stated in the consent and assent documents, and participants or guardians are able to opt out of this data sharing at any time. Opting out of NDA data sharing does not in any way affect trial eligibility.

#### Competing Interests

Dr Croarkin has received research grant support from Neuronetics Inc, NeoSync Inc, and Pfizer Inc. He has received in-kind support (equipment, supplies, and genotyping) for research studies from Assurex Health Inc, Neuronetics Inc, and MagVenture Inc. He has consulted for Engrail Therapeutics Inc, Myriad Neuroscience, Procter & Gamble, and Sunovion. Dr Vande Voort has received in-kind support (supplies and genotyping) from Assurex Health Inc. The other authors declare that they have no competing interests.

## Funding

This project is supported by Grant Number 1 R01 MH124655-01 from the NIMH, National Institutes of Health (NIH), and by Grant Number UL1 TR000135 from the National Center for Advancing Translational Sciences (NCATS). The content of this protocol and publication is solely the responsibility of the authors and does not necessarily represent the official views of the NIH.

## Authors’ Contributions

P.E.C. is the principal investigator, conceived the study concept, and spearheaded protocol development. P.A.N., M.J.Z., J.L.V., J.M.L., J.S., and P.E.C. designed the study aims, hypotheses, and statistical analyses. D.Y., C.O., and J.F.G. drafted the protocol, provided additional intellectual input, and made substantial revisions. All authors read and approved the final manuscript.

## Authors’ Information

See title page.

## Abbreviations

AE: adverse event
aTBS: accelerated theta burst stimulation
CDRS-R: Children’s Depression Rating Scale Revised
CRF: case report form
C-SSRS: Columbia–Suicide Severity Rating Scale
DPFC: dorsolateral prefrontal cortex
DSMB: Data and Safety Monitoring Board
EEG: electroencephalography
EMG: electromyography
FDA: US Food and Drug Administration
GABA: γ-aminobutyric acid
GEE: generalized estimating equation
HIPAA: Health Insurance Portability and Accountability Act
IDE: investigational device exemption
IRB: institutional review board
LICI: long-interval intracortical inhibition
LICI-100: LICI at an interstimulus interval of 100 ms
MDD: major depressive disorder
MINI-KID: Mini-International Neuropsychiatric Interview for Children and Adolescents
MNAR: missing not at random
MT: motor threshold
N45: 45 ms after stimulation
N100: 100 ms after stimulation
NDA: US National Institute of Mental Health Data Archive
NIH: US National Institutes of Health
NIMH: US National Institute of Mental Health
OR: odds ratio
PHI: protected health information
PI: principal investigator
SAE: serious adverse event
TBS: theta burst stimulation
TMS: transcranial magnetic stimulation
UADE: unanticipated adverse device effect

## Box.

### Inclusion and Exclusion Criteria

Inclusion criteria:

- Age 12-18 y
- Willingness and ability to give informed consent if aged 18 y or if <18 y, to assent (with consent given by a parent or legal guardian)
- Diagnosis of major depressive disorder (MDD) based on the criteria of the *Diagnostic and Statistical Manual for Mental Disorders* (Fifth Edition) for all participants and at screening the Mini-International Neuropsychiatric Interview (MINI) for Children and Adolescents (MINI-KID) for participants aged 12-17 or the MINI for participants who are 18
- Current episode of MDD lasting at least 4 weeks but <3 y
- Depressive symptom severity as shown by a total Children’s Depression Rating Scale Revised (CDRS-R) score ≥40
- Suicidal ideation score ≥3 on item 13 of the CDRS-R
- Minimum score of 1 (“wish to be dead”) on the Columbia–Suicide Severity Rating Scale (C-SSRS) severity of ideation subscale
- No demonstrated improvement by 25% or more in depressive symptom severity between screening evaluation and baseline
- Eligibility for transcranial magnetic stimulation (TMS) based on safety criteria
- Willingness to use a medically acceptable form of birth control during the 10-d treatment course if female and of childbearing potential

**Exclusion criteria:**

- Diagnosis of a psychotic disorder, bipolar disorder, anorexia nervosa, bulimia nervosa, or substance use disorder (except caffeine and tobacco) within the past year
- Lifetime diagnosis of a psychotic disorder, including schizophrenia, MDD with psychotic features, and bipolar disorder with psychotic features, confirmed by a research screening interview
- IQ <70^a^
- Positive urine drug screen at baseline
- History of epilepsy or unexplained seizures or any other neurologic condition that includes a history of seizures
- Family history of epilepsy
- Use of medications that lower the seizure threshold (eg, neuroleptic agents and tricyclic antidepressants)
- History of any treatment with electroconvulsive therapy or TMS
- Use of any investigational drug within 4 wk of baseline
- Prior brain surgery
- Risk of increased intracranial pressure (eg, brain tumor) or a history of increased intracranial pressure
- Lifetime history of head trauma with loss of consciousness for >5 min
- Any true-positive findings on the Transcranial Magnetic Stimulation Safety Screen
- Pregnancy or nursing
- Conductive, ferromagnetic, or other magnetic-sensitive metals in the head within 30 cm of the treatment coil (excluding the mouth) that cannot be safely removed (eg, cochlear implants, vagus nerve stimulators, deep brain stimulators, implanted electrodes/stimulators, aneurysm clips or coils, stents, bullet fragments, jewelry, and hair barrettes)
- Any implanted stimulator or other implant controlled by physiologic signals, including a vagus nerve stimulator, spinal cord stimulator, peripheral nerve stimulator, defibrillator, or pacemaker
- Implanted medication pump
- Cerebrovascular disease, cerebral aneurysm, dementia, movement disorder, or repetitive or severe head trauma, or primary or secondary tumors in the central nervous system
- History of severe headaches within the previous year
- Vascular, traumatic, tumoral, infectious, or metabolic lesions of the brain, even without a history of seizure or without anticonvulsant medication
- An unstable medical illness
- Improvement in depressive symptom severity between screening and baseline by ≥25%
- Inability to adhere to the protocol

## Legend

**Figure.**
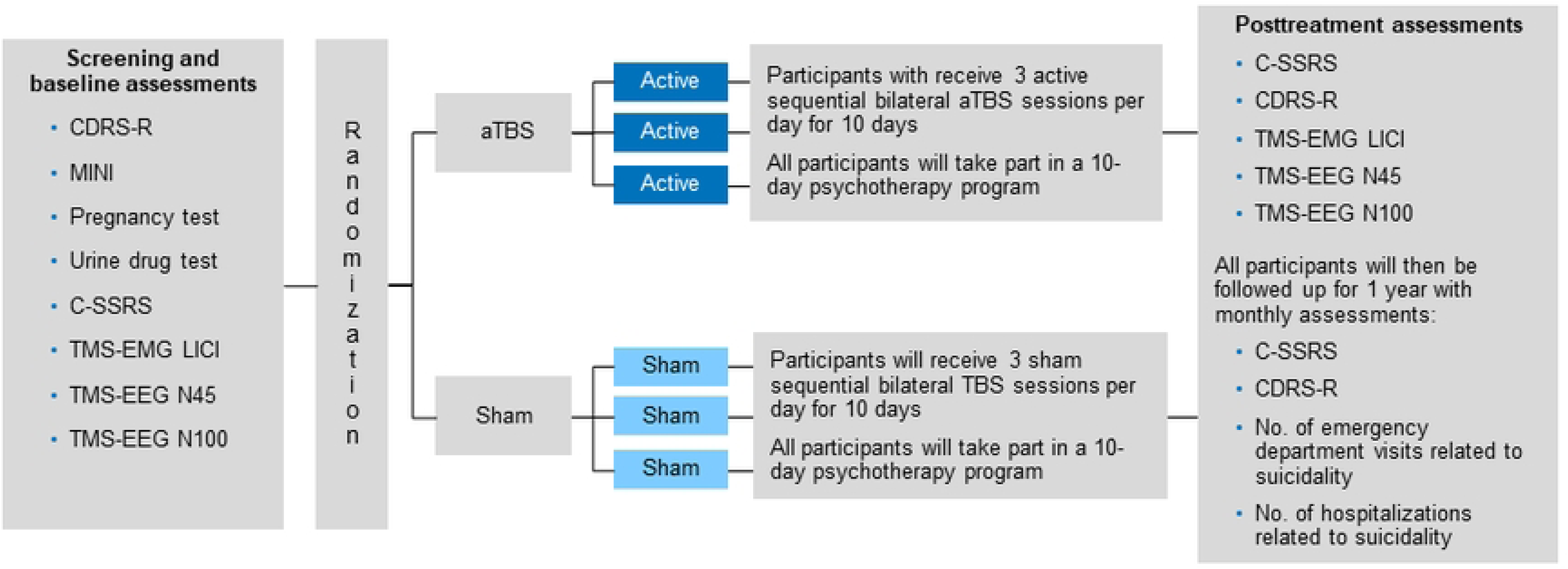
Study Schema. aTBS, accelerated theta burst stimulation; CDRS-R, Children’s Depression Rating Scale Revised; C-SSRS, Columbia–Suicide Severity Rating Scale; LICI, long-interval intracortical inhibition; MINI, Mini-International Neuropsychiatric Interview; N45, TMS-EEG evoked potential at 45 milliseconds; N100, TMS-EEG evoked potential at 100 milliseconds; TBS, theta burst stimulation; TMS-EEG, transcranial magnetic stimulation and electroencephalography; TMS-EMG, transcranial magnetic stimulation and electromyography.

## Footnotes

a ^a^ If a clinical concern exists about IQ, participants will be psychometrically assessed with the Slosson Intelligence Test–Revised, 3rd edition (Slosson RL. Slosson Intelligence Test-Revised. 3rd ed. Revised by Nicholson CL, Hibpshman TL, Supplementary Manual by Larson S. East Aurora, NY: Slosson Educational Publications Inc).

## Notes

### Clinical Trial

ClinicalTrials.gov (ID: NCT04701840) https://clinicaltrials.gov/ct2/show/NCT04502758?term=NCT04701840&draw=2&rank=1

### Funding Statement

This project is supported by Grant Number 1 R01 MH124655-01 from the NIMH, National Institutes of Health (NIH) (awarded to PEC), and by Grant Number UL1 TR000135 from the National Center for Advancing Translational Sciences (NCATS) (awarded to PEC). The funders had and will not have a role in study design, data collection and analysis, decision to publish, or preparation of the manuscript.The content of this protocol and publication is solely the responsibility of the authors and does not necessarily represent the official views of the NIH.

### Author Declarations

This protocol is approved by the Mayo Clinic IRB (20-009630), NIMH DSMB, and FDA Investigational device exemption (IDE) Number: G200220

